# Feasibility and sociopsychological impact of video consultations in medical oncology - a randomized controlled open label trial

**DOI:** 10.1101/2020.07.10.20150052

**Authors:** Thomas Walle, Erkin Erdal, Leon Mühlsteffen, Hans Martin Singh, Editha Gnutzmann, Barbara Grün, Helene Hofmann, Alexandra Ivanova, Bruno Köhler, Felix Korell, Athanasios Mavratzas, Andreas Mock, Constantin Frederic Pixberg, David Schult, Helen Starke, Niels Steinebrunner, Lena Woydack, Andreas Schneeweiss, Mareike Dietrich, Dirk Jäger, Johannes Krisam, Jakob N. Kather, Eva C. Winkler

**Affiliations:** Department of Medical Oncology, National Center for Tumor Diseases (NCT), Heidelberg, Germany; Department of Medical Oncology, University Hospital Heidelberg, Heidelberg, Germany; Clinical Cooperation Unit Virotherapy, German Cancer Research Center (DKFZ), Heidelberg, Germany; German Cancer Consortium (DKTK), Heidelberg, Germany; Department of Medicine II, University Hospital Hamburg Eppendorf, Hamburg, Germany; Minxli AG, München, Germany; Department of Hematology, University Hospital Heidelberg, Heidelberg, Germany; Institute of Medical Biometry and Informatics, University Hospital Heidelberg, Heidelberg, Germany; RWTH University Hospital Aachen, Department of Medicine III, Aachen, Germany

## Abstract

**Background:** Mobile phone video call applications generally did not undergo testing in randomized controlled clinical trials prior to their implementation in patient care regarding the rate of successful patient visits and impact on the physician-patient relationship.

**Methods:** The NCT MOBILE trial was a monocentric open-label randomized controlled clinical trial of patients with solid tumors undergoing systemic cancer therapy with need of a follow-up visit with their consulting physician at outpatient clinics. 66 patients were 1:1 randomized to receive either a standard in-person follow-up visit at outpatient clinics or a video call *via* a mobile phone application. The primary outcome was feasibility defined as the number of successful appointments at the first follow up visit. Secondary outcomes included success rate of further video calls, time spent by patient and physician, patient satisfaction, and quality of physician-patient relationship.

**Findings:** Success rate of the first follow up visit in the intention-to-treat cohort was 87.8% for in-person visits and 78.7% for video calls (p=0.51, RR=0.88-1.43 95%CI). The most common reasons for failure were software incompatibility (12%) in the video call and no-show (6%) in the in-person visit arm. The success rate for further video visits was 91.6% (11 of 12 calls). Standardized patient questionnaires showed significantly decreased total time spent and less direct costs for patients (Δ95 to 246min 95%CI, Δ4.8 to 23.9€ 95%CI) and comparable time spent for physicians in the video call arm (Δ-6.4 to 5.4min, 95%CI). Doctor-patient relationship quality mean scores assessed by the validated standardized “questionnaire on quality of physician-patient interaction”(QQPPI) were higher in the video call arm (video call/in-person = 1.12 fold, p=0.02).

**Interpretation:** Follow-up visits with the tested mobile phone video call application were feasible but software compatibility should be critically evaluated.

**Trial registration:** Retrospectively registered in the German Clinical Trials Register DRKS00015788, 26^th^ October 2018

## Background

Patients with solid tumors frequently undergo systemic cancer therapy for many months, especially in the metastatic setting. To monitor and treat adverse events and infections these patients frequently need to consult with their medical oncologist. Commutes to outpatient clinics of specialized comprehensive cancer centers can be long and strenuous for this fragile patient population. A retrospective analysis recently suggested that palliative systemic cancer therapy including commutes to clinics accounted for approximately 10% of the survival time awake remaining to pancreatic cancer patients with distant metastases (1).

Telemedicine applications can facilitate patient access to specialized healthcare from remote and may therefore be ideally suited for the medical oncology setting. With the recent SARS-CoV-2 pandemic the need for remote healthcare has risen substantially requiring thoroughly tested telemedicine applications.

Functions of telemedicine applications include scheduling appointments to consult with healthcare professionals *via* encrypted video call, monitoring of symptoms or treatment adherence as well as patient education (2, 3). Telemedicine applications are now increasingly offered for common smartphone operating systems to make access to healthcare even more convenient and independent of desktop computer access. Healthcare providers and insurers have implemented mobile phone applications including video call applications to facilitate their patients’ access to healthcare.

Although there is evidence that these applications may reduce costs without negatively affecting clinical outcomes (3, 4), few commercial applications underwent testing in randomized controlled trials prior to their clinical implementation (5, 6). In vulnerable oncology patients undergoing systemic cancer therapy it therefore remains unclear (1) how robust these applications are with regards to their failure rate and (2) how telemedicine applications affect communication strategies of physicians such as shared-decision making and the resulting patient-physician relationship.

The primary objective of the NCT MOBILE trial was to assess feasibility defined as the failure rate of video consultations as compared to in-person visits in patients with solid tumors undergoing systemic cancer therapy who required a follow-up appointment. By using standardized and validated questionnaires we also assessed patient satisfaction, the economic impact and the sociopsychological effects of these video consultations.

## Methods

### Study design

The NCT MOBILE trial was a randomized controlled open-label clinical trial at the National Center for Tumor Diseases (NCT) in Heidelberg, Germany. Patients with solid tumors (ICD-10 2016, C00-C97) undergoing systemic cancer therapy and requiring a follow-up visit in 2-14 days’ time were recruited by medical oncologists at NCT outpatient clinics. Patients were 1:1 randomized to receive their follow-up appointment at outpatient clinics (in-person) or *via* a dedicated smartphone application in German language “Minxli – Arzt via Video Chat”(https://www.minxli.com/). The outcome of the appointment was documented by the treating physician in the case report form (CRF, 10.5281/zenodo.3902837) and by the patient in questionnaire “Q1”(10.5281/zenodo.3902837). Patients in the video call group were eligible to schedule further video calls *via* the mobile phone application. After completion of oncological therapy or 6 months after randomization patients in the video call group were asked to fill out questionnaire “Q2”. Ethical approval was granted by the ethics commission of the medical faculty at Heidelberg University (S-090/2017). The trial was registered in the German Clinical Trial Register (DRKS00015788) where the original study protocol and ethics committee approval can be found (https://www.drks.de/drks_web/navigate.do?navigationId=trial.HTML&TRIAL_ID=DRKS00015788). No changes to the study protocol were made after trial initiation. Patients were recruited from 29^th^ November 2017 (first patient in) until 7^th^ October 2019 (last patient in), follow up period was 6 months for all patients with the last follow-up period ending on 7^th^ April 2020.

### Participants

Patients were eligible for inclusion in the trial if they were 18 years or older, had a performance status of Eastern Cooperative Oncology Group (ECOG) 0-2, owned a compatible smartphone with Android (Google LLC, Mountain View, CA, U.S.A.) or iOS (Apple Inc., Cupertino, CA, U.S.A.) operating system and were comfortable using it and agreed to the “Minxli – Arzt via Video Chat”terms and conditions. Patients not proficient in the German language or patients with severe visual or auditory impairments were excluded from participation in the trial. All patients provided written informed consent.

### Randomization procedure

Sequentially labeled sealed opaque envelopes were prepared by the study’s statistician (J.K.) who remained blinded to treatment allocation until all treatments had been allocated. Block randomization was used, and everybody was blinded to block length except the statistician. For each patient one envelope was opened by T.W., J.N.K. or L.M. or E.G. after the respective patient had provided informed written consent. Treatment allocation was conveyed to the patient and treating physician in an open-label design.

### Mobile phone application

Patients were instructed to download and install the smartphone application “Minxli – Arzt via Video Chat”(version 1.3.1) from the Google Play Store (Google LLC, Mountain View, California, U.S.A.) or the application “Minxli – Arzt via Video Chat”(version 1.2.8) from the Apple App Store (Apple Inc., Cupertino, California, U.S.A.). The application was provided in German language and compatible with the operating systems iOS v10 or higher (Apple Inc., Cupertino, California, U.S.A.) and Android v4.4 or higher (Google LLC, Mountain View, California, U.S.A.). Key features of the mobile application included scheduling encrypted video calls with verified physicians, a chat function with options to upload pictures, which was only available when a valid appointment had been scheduled by the patient and confirmed by the physician and a medication plan management function. Video calls were initialized by the physicians and were only possible after an appointment had been scheduled by the patient and confirmed by the physician, thereby preventing patients from calling their physicians at unscheduled times. In this study, verification of physician identity was guaranteed by the study lead. Video call data was transmitted using end-to-end encryption. All protected health information was encrypted in transit and stored on Amazon Web Services Simple Storage System (Amazon.com Inc, Seattle, Washington, U.S.A.) servers located in Frankfurt, Germany.

### Outcomes

The primary outcome was feasibility defined as the number of successfully completed appointments in the video call and in-person consultation group. A successful appointment was defined as a medical consultation between patient and physician that was unanimously finished and was not cancelled because of technical issues or other problems.

Secondary outcomes included patient satisfaction, content of the appointments, quality of the patient-physician relationship and cost- and time-efficiency during the first appointment as assessed in questionnaire “Q1”which had to be filled out directly after the appointment to avoid recall bias. Further information about the time spent for the appointment by the treating physician and content of the appointment was assessed in the CRF (10.5281/zenodo.3902837).

Total time spent was calculated according to the following formula. T_total_ = 2*T_travel_+ T_wait_, with T_total_: total time spent, T_travel_: time spent for one-way commute from home to NCT outpatient clinics, T_wait_: total waiting time spent at NCT outpatient clinics. Direct costs were reported by patients and indirect costs were calculated according to the following formula. costs_indirect_ = T_work_ * S, with costs_indirect_: indirect costs, T_work_: time absent from work due to the follow-up appointment as reported by the patient, S: average salary in the state of Baden-Württemberg, Germany of 24€/h (Statistisches Landesamt Baden-Württemberg, press release 158/2018, Stuttgart, Germany 12.7.2018, URI: http://www.statistikbw.de/Presse/Pressemitteilungen/2018158). T_work_ was set to 0 for all patients without employment, on sick-leave and retired patients.

After completion of the study, the general experience with the mobile phone application was assessed using questionnaire “Q2”. Age, gender, post code, oncological main diagnosis, UICC stage and time of initial oncological diagnosis were retrieved from NCT’s electronic medical documentation system. ECOG and therapy scheme were retrieved from the case report forms. Straight-line-distance from the supplied postal code to the hospital was calculated using Google Maps (Google LLC, Google LLC, Mountain View, CA, U.S.A.).

### Questionnaire Design

Our interdisciplinary research team (social scientist, medical oncologist, medical ethicist, IT-specialist) developed the questionnaires “Q1”(10.5281/zenodo.3902837) and “Q2”(10.5281/zenodo.3902837) in German language based on a validated German instrument for the patient-physician interaction and self-developed questions adapted from prior assessments of telemedicine outcomes (7).

Hence, “Q1”consisted of two sections of self-developed questions and one validated instrument. Section 1 (13 items) assessed the general experience, time and money spent for the first video call or in-person appointment, section 2 (7 items) reported the content of appointment (physical and instrumental examinations, prescriptions etc.) using five-point Likert scale to different statements. Section 1&2 also included open questions to which patients could respond in free text. Section 3 (13 items) assessed the physician-patient relationship using the validated questionnaire on quality of physician-patient interaction (QQPPI) in German language (8-10). To calculate the QQPPI score Likert levels of all items were summed per patient with a high score indicating high and a low score indicating low satisfaction of the interaction with the physician, respectively. Part 3 was only calculated for patients who completed all 13 questions because the QQPPI score lacks validation for incomplete questionnaires.

“Q2”assessed the desire to repeat the appointment, technical difficulties, and the number of appointments in total as remembered by the patient at the end of oncological therapy or 6 months after treatment initiation. “Q2”consisted out of 10 self-developed items, of which 5 were open questions to be answered in free-text and 5 were five level Likert scale items.

All questionnaires were sent back to the study lead in a pseudonymized format. To report this study most transparently, we provide all original questionnaires as well as their translations from the German to the English language under a Creative Commons Attribution 4.0 International License (http://doi.org/10.5281/zenodo.3902837)

### Statistical analysis

The primary outcome parameter was compared between patients in the video call and in-person visit group using Fisher’s exact test (two-sided). Patients who obviously evaluated the wrong appointment in questionnaire Q1 were excluded from analysis (n=3, e.g. video call patients indicating journey from home). Likert-scale scores were compared using Mann-Whitney-U tests (two-sided), spending in € and time-spent were compared using unpaired t-tests (two-sided). Multiple comparisons were accounted for using the Benjamini-Hochberg method. Statistically significant comparisons are indicated with an asterisk. No statistical sample size estimation was performed for this trial. P values and confidence intervals were calculated using GraphPad Prism v8.4.2 for Windows 10 (GraphPad Software LLC). Benjamini-Hochberg correction was calculated using the p.adjust function in the R base package (R Studio 1.2.5003).

## Results

Between 29.11.2017 and 10.07.2019 we screened 306 patients as potentially eligible **(Figure 1)**. Of these, 43 patients (14%) were deemed ineligible prior to randomization due to insufficient proficiency in the German language (n=22), ECOG 3 or 4 (n=8), the need of in-house diagnostics (n=6), incompliance (n=4) or severe auditory or cognitive impairments (n=3). Of the remaining patients, 29 (9%) objected to randomization because they solely preferred video calls and 105 (34%) were unwilling to participate due to unspecified reasons. Another 55 patients (17.9%) were not adept at using or not owning a smartphone a priori compatible with the application (iOS v10 or higher, Android v4.4 or higher) and 8 patients (2%) had concerns regarding the safety of the video call.

**Figure 1.**
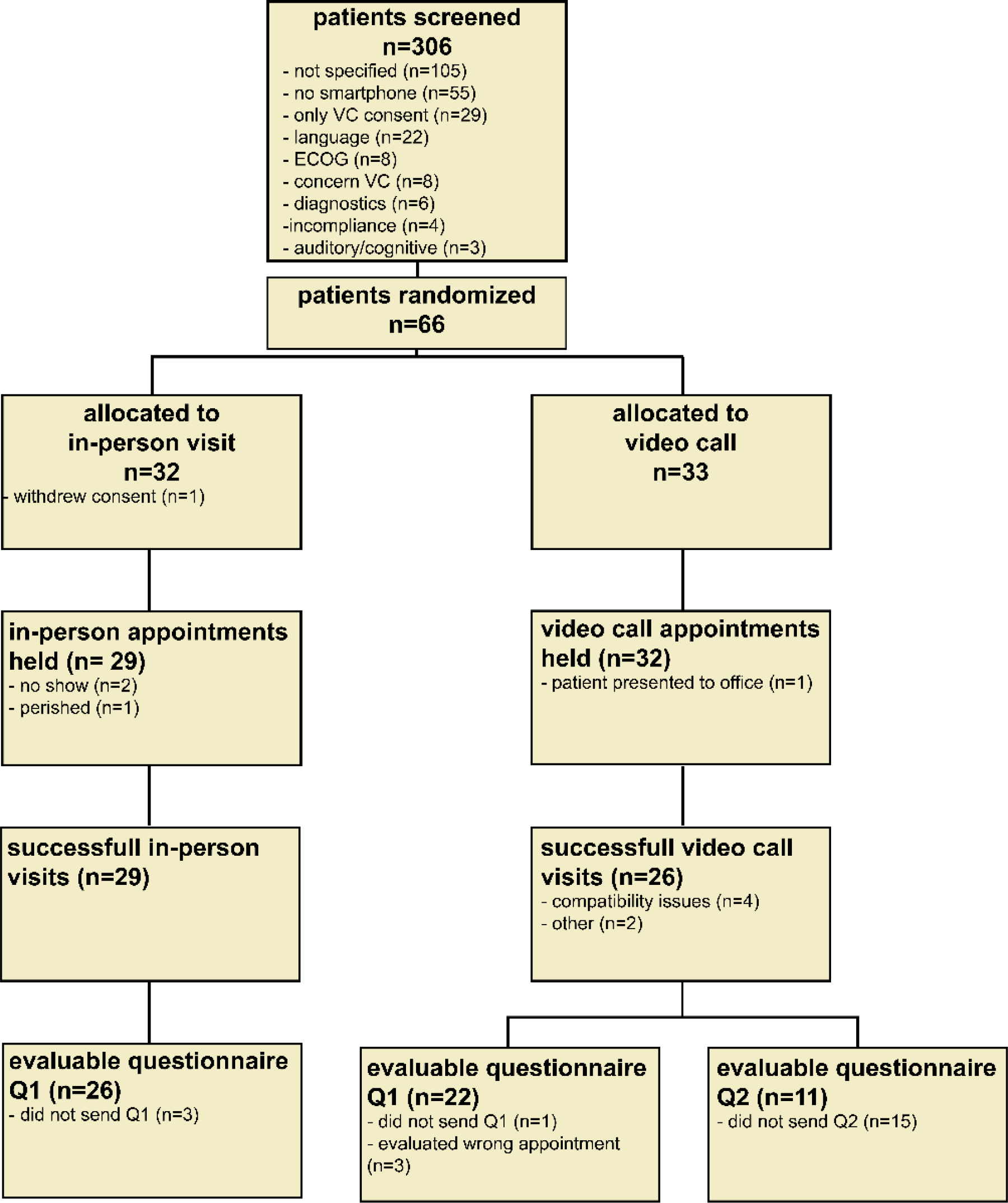
CONSORT flowchart. Consolidated Standards of Reporting Trials (CONSORT) flow chart indicating screening, randomization, and data completeness. Reasons for exclusion/failure are highlighted with bullet points. ECOG: Eastern Co-operative Oncology Group performance status, no smartphone: patients not adept at using or not owning a smartphone, VC: video call, Q1: questionnaire Q1, Q2: questionnaire Q2.

This resulted in a total of 66 patients who were 1:1 randomized to the video call (n=33) and in-person visit (n=33) arm **(Figure 1)**. One patient in the in-person visit arm withdrew consent after randomization resulting in a total of 32 evaluable patients. The cohort included patients with a range of different tumor types and therapies including cytotoxic chemo-, targeted and immunotherapy in either palliative or (neo)adjuvant intention **(Table 1)**. Patients in the video call and in-person visit cohorts showed representative age distribution for medical oncology patients. ECOG performance status and distance to the hospital were similar in both groups. However, we observed a higher number of female patients with breast cancer in the video call (n=10) as compare to the in-person visit group (n=2), resulting in a higher number of Union for International Cancer Control (UICC) stage 1 patients (n=6 vs n=1) in the video call arm.

**Table 1.** Patient baseline characteristics. Table indicating patient characteristics at baseline. anti-PD-1/PD-L1: pembrolizumab, nivolumab, avelumab, atezolizumab, durvalumab; BLCA: bladder urothelial carcinoma; BRCA: breast cancer; CESC: cervical squamous cell carcinoma; CisBev: cisplatin + bevacizumab; CRC: colorectal adenocarcinoma; CUP: carcinoma of unknown primary; DC: doxorubicin + cyclophosphamide; Doce: docetaxel monotherapy; EC: epirubicin+cyclophosphamide, ESCA: esophageal carcinoma; FOLFIRI (+/- mab): FOLFIRI, FOLFIRI+aflibercept, FOLFIRI+bevacizumab, FOLFOX/FLO/CapOx +/- mab: FOLFOX, FOLFOX/FLO/CapOx + cetuximab/panitumumab/bevacizumab/aflibercept, HCC: liver hepatocellular carcinoma; HNSCC: head and neck squamous cell carcinoma; NET: neuroendocrine tumor; OV: ovarian adenocarcinoma; no.: number; platinum triplet: FOLFIRINOX or FOLFOXIRI + bevacizumab or cisplatin + paclitaxel + gemcitabine or FLOT; PCA: prostate adenocarcinoma; PDAC: pancreatic adenocarcinoma; platin+taxane: carboplatin + paclitaxel, TCbHP schema (docetaxel, carboplatin, trastuzumab, pertuzumab), cabazitaxel+carboplatin; RCC: clear cell renal cell carcinoma; SARC: soft tissue sarcoma; GC: gastric adenocarcinoma; UCEC: uterine corpus endometrium cancer; VUL: vulvar squamous cell carcinoma; yr: years.

| GENERAL | In-person (n=32) | Video call (n=33) |
| --- | --- | --- |
| Patient number | 32 | 33 |
| Age – yr |  |  |
| Median | 59.5 | 54 |
| Range | 29-72 | 22-74 |
| Sex – no. (%) |  |  |
| Female | 13 (40.6) | 17 (51.5) |
| Male | 19 (59.3) | 16 (48.4) |
| ECOG performance status score – no. (%) |  |  |
| 0 | 14 (43.7) | 17 (51.5) |
| 1 | 15 (46.8) | 14 (42.4) |
| 2 | 3 (9.3) | 2 (6.0) |
| UICC stage – no. (%) |  |  |
| 1 | 1 (3.1) | 6 (18.1) |
| 2 | 2 (6.2) | 3 (9.0) |
| 3 | 2 (6.2) | 6 (18.1) |
| 4 | 27 (84.3) | 18 (54.5) |
| Tumor type – no. (%) | CRC 9 (28.1)<br>PDAC: 7 (21.8)<br>PCa: 3 (9.3)<br>BRCA: 2 (6.2)<br>HNSCC: 2 (6.2)<br>CUP: 1 (3.1)<br>ESCA: 1 (3.1)<br>RCC: 1 (3.1)<br>NET: 1 (3.1)<br>OV: 1 (3.1)<br>SARC: 1 (3.1)<br>GC: 1 (3.1)<br>UCEC: 1 (3.1)<br>VUL: 1 (3.1) | BRCA: 10 (30.3)<br>CRC: 5 (15.1)<br>PDAC: 4 (12.1)<br>PCa: 3 (9.0)<br>GC: 3 (9.0)<br>BLCA: 2 (6.0)<br>NET: 2 (6.0)<br>CESC: 1 (3.0)<br>ESCA: 1 (3.0)<br>HNSCC: 1 (3.0)<br>HCC: 1 (3.0) |
| Therapy scheme – no (%) | Platinum triplet (+/- mab): 8(25.0)<br>FOLFOX/FLO/CapOx (+/- mab):5(15.6)<br>anti-PD-1/PD-L1: 4(12.5)<br>FOLFIRI (+/- mab): 4(12.5)<br>Platin+Taxane(+/- mab): 4(12.5)<br>CisTopo (+/- mab): 2(6.25)<br>Docetaxel: 2(6.25)<br>Gem+nabPac: 1(3.125)<br>EC/DC: 1(3.125)<br>INN-doxorubicin: 1(3.125) | EC/DC: 8(24.2)<br>Platinum triplet (+/- mab): 6(18.1)<br>FOLFOX/FLO/CapOx (+/- mab):5(15.1)<br>anti-PD-1/PD-L1: 4(12.1)<br>FOLFIRI (+/- mab): 4(12.1)<br>Doce: 2(6.0)<br>CisBev: 1(3.0)<br>Epirubicin: 1(3.0)<br>Platin+Taxane(+/- mab): 1(3.0)<br>Sunitinib/Lanreotid: 1(3.0) |
| Linear distance to hospital – km (sd) | 34.1 (38.7) | 28.4 (20.5) |

The first appointment took place as scheduled in 90% (n=29) of patients of the in-person visit and 96% (n=32) of patients in the video call arm **(Figure 1)**. In the in-person visit arm, 2 patients did not present to outpatient clinics and could not be contacted by any means. One patient died before the scheduled appointment. In the video call, arm one patient erroneously presented to outpatient clinics.

The remaining 29 patients in the in-person visit arm completed the appointment successfully. In the video call arm, 81.2% (n=26) of patients completed their appointments *via* mobile phone application successfully. 18.75% (n=6) of patients experienced technical difficulties resulting in premature termination of the video call appointment. These patients could be contacted by phone and did not have to present to outpatient clinics. The most common reasons for failure were compatibility issues (66.6%, n=4), followed by unstable internet connection (16.6%, n=1) and problems with the appointment scheduling function of the mobile phone application (16.6%, n=1). In summary, this resulted in a success rate of 87.8% for in-person visits and 78.7% for video calls in the intention-to-treat cohort, which was not significantly different (p=0.51, RR=0.88-1.43 95%CI) **(Figure 2A)**. The success rate for further video calls after the first appointment was 91.6% **(Figure 2B)**. Technical difficulties were not associated with age (failure: median=53 years, all video calls: median=54 years) but we detected a non-significant difference in gender with more males experiencing technical difficulties (failure: 80% male, success: 42% male, p=0.65).

**Figure 2.**
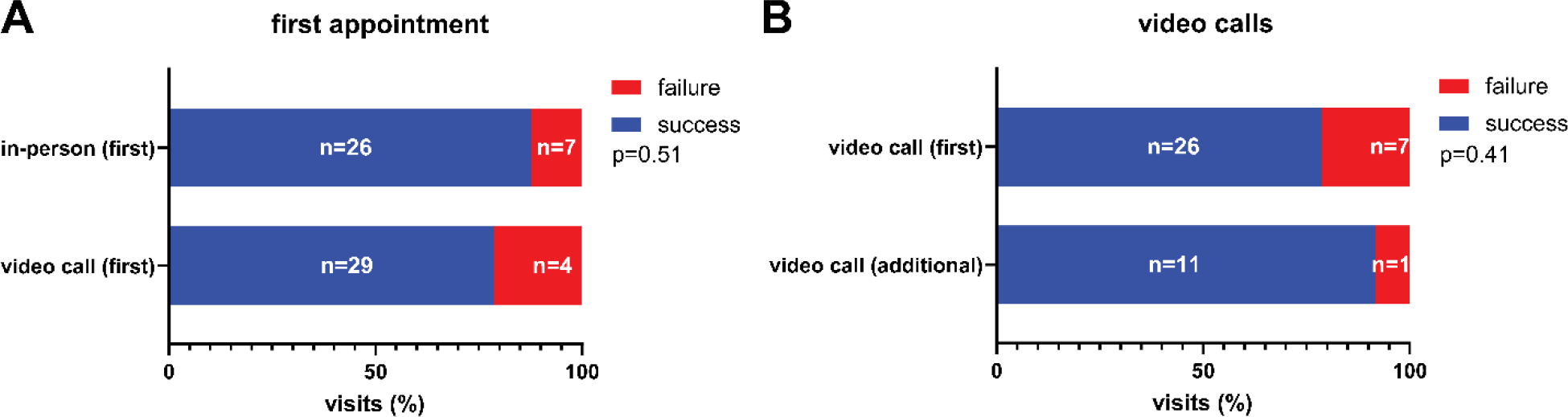
Success rate of video call and in-person visits. Stacked bar graphs indicating success rates of patients in the in-person visit and video call arms. **(A)** Success rates at first scheduled appointment in the in-person and video call arms. **(B)** Success rates for video call appointments at the first scheduled and at any additional appointments. P-values were calculated using Fisher’s exact test.

Patients who successfully completed in-person and video visits were asked to fill out a questionnaire (Q1) directly after the appointment to avoid recall bias. We received 26 evaluable Q1 questionnaires in the in-person and 22 in the video call group **(Figure 1)**. We assessed the content of the video visits including physical and instrumental examinations, administered treatments, prescriptions, referrals, and scheduling of additional follow-up appointments. These contents differed from in-person appointments although these differences did not reach statistical significance **(Figure 3A)**. Physical examinations were performed in 30% of in-person as compared to 9% of video calls. Physicians filled out prescriptions in 50% of the in-person as compared to 9% of the video calls. Moreover, medication was directly administered in 7% of the in-person appointments. Additional appointments had to be scheduled in 4.5% of the video calls but not in the in-person group. Physicians referred patients to other health care professionals in 11.5% of the in-person but only in 4.5% of the video call appointments.

**Figure 3.**
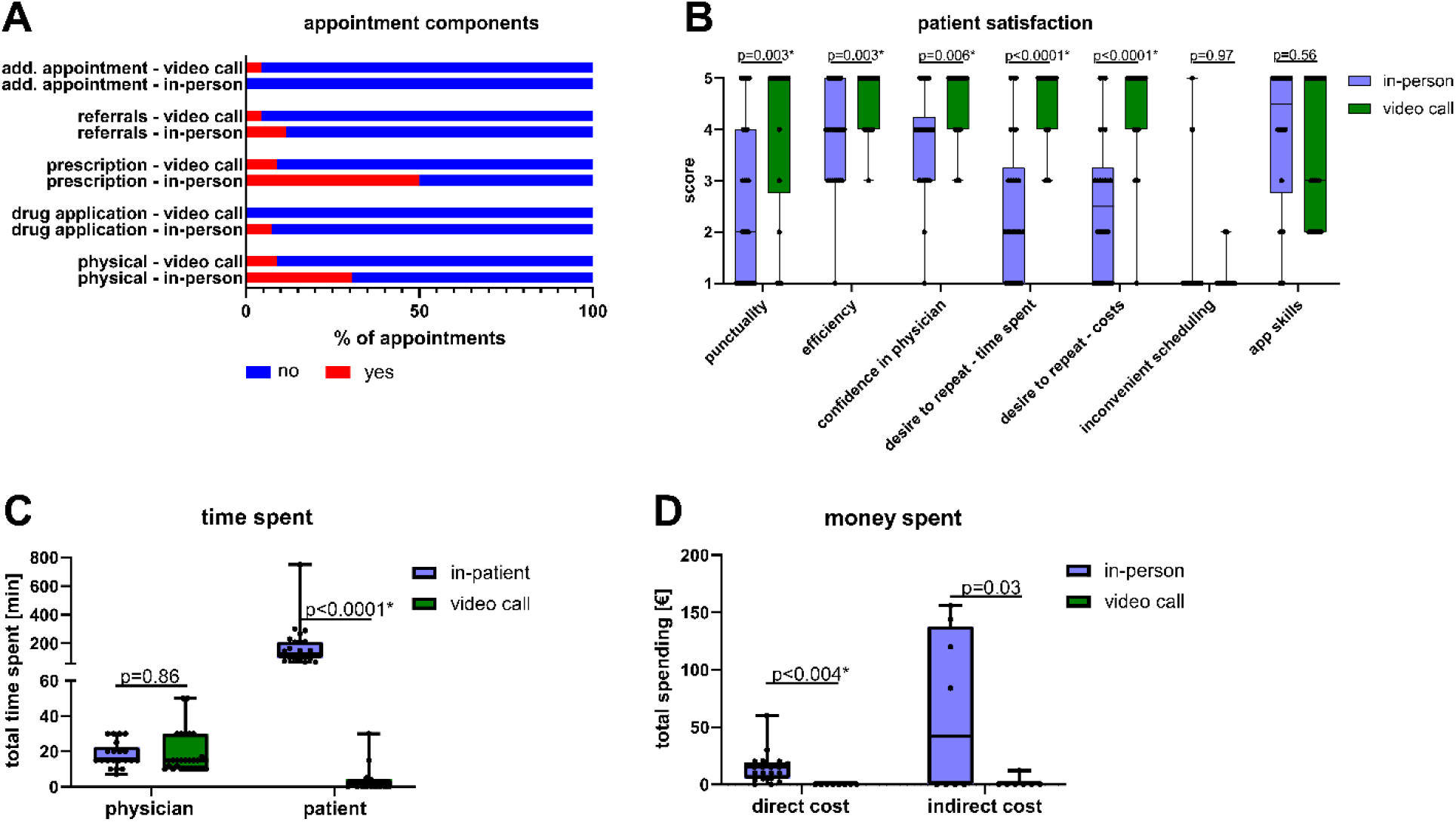
Appointment characteristics and patient satisfaction. Components of the appointment, patient satisfaction, time, and cost were assessed for the first scheduled appointment. **(A)** Stacked bar graphs indicating characteristics of the first appointment in the in-person and video call arms. **(B)** Box plots indicating different dimensions of patient satisfaction and the desire to repeat the appointment in the in-person (n=26) or video call (n=22) group. P-values were calculated using Mann-Whitney-U tests (two-sided). **(A-B)** Indicated are descriptive titles for the items, the complete items can be found in Figures S3-4. **(C)** Box plots indicating total time spent for physicians (n=47) and patients (n=39). P-values were calculated using unpaired t-tests (two-sided). **(D)** Box plots indicating total direct (n=29) and indirect costs (n=15) for patients in the in-person and video call arm. P-values were calculated using unpaired t-tests (two-sided). **(B-D)** Multiple comparisons were accounted for using the Benjamini-Hochberg method. Statistically significant comparisons are indicated with an asterisk (q<0.05). Boxes indicate interquartile range, bars indicate median and whiskers range.

Patients indicated higher overall satisfaction in the video call group **(Figure 3B)**. Patients ranked confidence in their physician (p=0.006), efficiency (p=0.003) and punctuality (p=0.003) higher in the video call group as compared to the in-person appointment **(Figure 3B)**. Accordingly, patients in the video call group preferred the video call setting for future visits with respect to saving time (p<0.0001) and cost (p<0.0001) **(Figure 3B)**. Indeed, patients in the video call group saved 170min on average (p<0.0001, 95 to 246min 95%CI) and 14.37€ in direct costs (p<0.004, Δ4.8 to 23.9€ 95%CI) as compared to the in-person visit group **(Figure 3C-D)**. This positive assessment of the video consultations and desire to repeat them was maintained throughout the study period as assessed by the end of study questionnaire Q2 **(Table S1)**.

Patients in the video call group saved an average of 61.28€ (p=0.039, Δ3.3 to 119.2€ 95%CI) in indirect costs due to reduced absence at work **(Figure 3D)**. Time spent was comparable for physicians in the video call and in-person visit arm **(Figure 3C)** (Δ-6.4 to 5.4min, p=0.86).

We assessed physician-patient relationship quality using the validated standardized “Questionnaire on the Quality of Physician-Patient Interaction”(QQPPI) which is characterized by high reliability (Cronbach’s α.97) and uni-dimensionality and has been successfully applied to randomized clinical trials (8, 9). We found that QQPPI scores were higher in patients of the video call as compared to the in-person visit group **(Figure 4A)**. This effect was consistent across the different items of the questionnaire **(Figure 4B)** suggesting that the physician-patient relationship is not negatively affected but may be positively affected by video calls.

**Figure 4.**
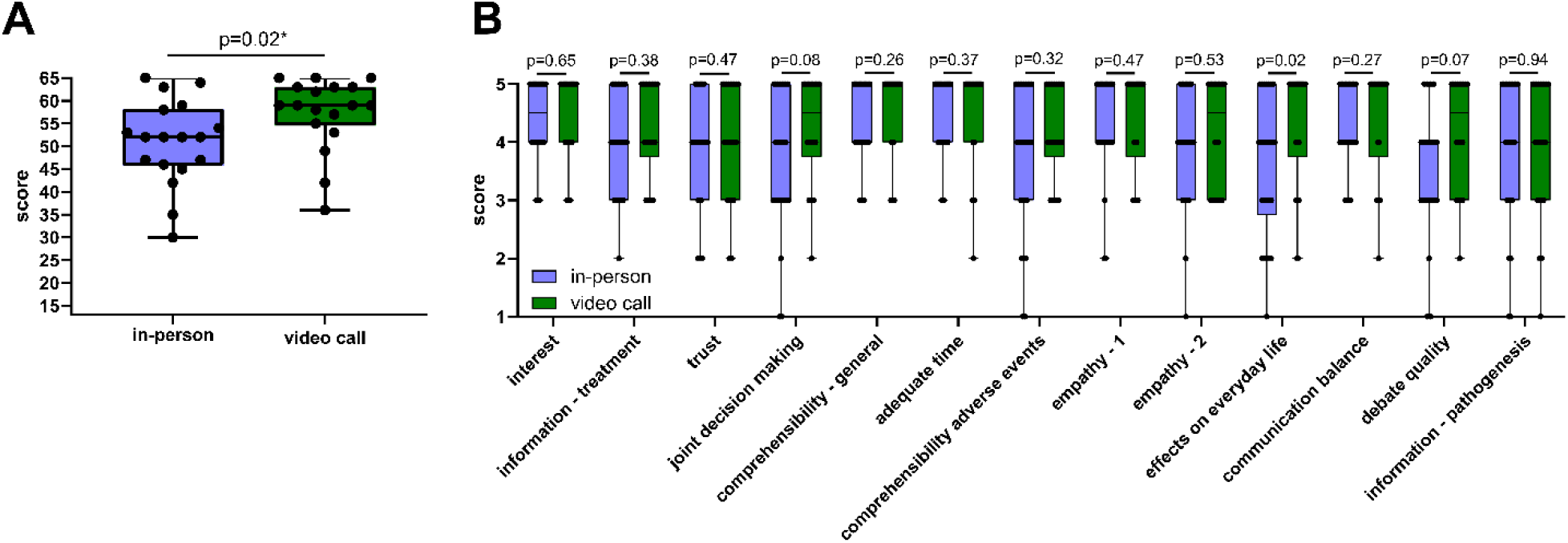
Physician-patient relationship assessment using the questionnaire on quality of physician-patient interaction. Physician-patient relationship after the first appointment was assessed using the questionnaire on quality of physician-patient interaction (QQPPI) questionnaire. **(A)** Box plots indicating QQPPI total score in the in-persons and video call groups. **(B)** Box plots indicating patient agreement with the individual 5 level Likert-scale items of the QQPPI questionnaire. Indicated are descriptive titles for the items, the complete items can be found in Figures S3-4. **(A-B)** P-values were calculated using Mann-Whitney-U tests. Multiple comparisons were accounted for using the Benjamini-Hochberg method. Asterisks indicate significant comparisons (q<0.05). Boxes indicate interquartile range, bars indicate median and whiskers range.

## Discussion

Here we present evidence that video call applications are generally feasible in medical oncology patients with no significant differences in success rate in the intention-to-treat analysis. However, this trial was not planned with a non-inferiority design prohibiting strong conclusions about comparability of failure rates. Despite low patient numbers, our cohort covered a broad range of tumor types and was representative for medical oncology patients regarding age and administered therapies.

Screening failures revealed non-availability of a smartphone or non-adeptness in its use as the most frequent reason for non-participation in the trial. Along these lines, poor availability and infrequent use of information and communication technology have been linked to other inequalities in non-digital health care access highlighting the risk that telemedicine may reinforce these social inequalities (11, 12). However, the net effect of telemedicine on social inequalities will ultimately depend on current non-digital health care access for underprivileged groups and the strength of digital illiteracy in these groups.

We observed different reasons for failure of the appointments in the video call and in-person visit group. In the video call group, we observed a relevant number of patients experiencing compatibility issues which could not be resolved by technical support. In further video calls which were only scheduled by patients without compatibility issues in the first visit, success rates were higher, although the number of further follow-up visits was low. In our study, none of the patients were at risk because of these technical difficulties. All video call patients who could not be contacted by the video call application were contacted by phone and did not have to present to outpatient clinics. However, it is conceivable that video call failures may lead to unscheduled visits in rare cases or alternatively may affect the physician’s ability to correctly assess a condition of a patient because this assessment also relies on visual inspection. Hence, compatibility of mobile phone applications should be critically investigated prior to their implementation.

In the in-person group, two patients did not present to outpatient-clinics and one patient withdrew consent after randomization suggesting that patients may skip appointments which they deem unnecessary due to the effort of presenting to outpatient clinics. Along these lines, 9% of all screened patients refused participation in the trial because they were only interested in the video call group highlighting the strong interest of patients to avoid supposedly dispensable commutes. In contrast, to the video call arm where all patients could be contacted, two no-show patients in the in-person visit arm could not be contacted by any means at the time of their scheduled appointment. In high-risk situations difficult to understand for the patient, such as profound neutropenia, ensuring compliance may be critical. In our cohort, compliance was better in the video call arm. Thus, our data suggest that compliance may be positively affected by use of the tested video call application.

Given the failure rates and reasons for failure of the appointment we observed in our video call and in-person visit groups, a nuanced evaluation of the setting used for an appointment may be adequate for clinical practice. For situations with digitally less-literate patients or risk factors for technical difficulties such as old operating systems and poor internet connection an in-person appointment should be scheduled. For digitally literate patients and a risk for non-compliance or no-show a video call seems more adequate based on the data presented. In situations where visiting outpatient clinics may present a considerable health hazard for patients such as during the recent SARS-CoV-2 pandemic, video calls may become a necessity for oncology patients. Along these lines, during the SARS-CoV-2 pandemic, cancer screenings and non-emergency in-person visits of new patients were frequently declined for the sake of epidemiological infectious disease control (13).

Similar to previous studies in other medical specialties, our patients were highly satisfied with their video call experience (3, 7, 14). Reasons for high patient satisfaction may be the observed cost and times savings, which were similar to previous reports (7, 14). For physicians, the application did not result in increased time expenditure. The application also prevented unscheduled consultations and enabled physicians to remain in full control of the initialization of the appointments. This may be an important factor to ensure work-life balance of physicians and increase acceptance of these applications in health-care workers. In our study, the patients’ perception of the patient-physician relationship was more positive in the video call group as compared to the in-person visit arm. This outcome was unexpected given frequent concerns about the importance of physical interactions voiced by health care professionals (15). Importantly, the physician-patient relationship is bidirectional, and we only assessed the patients’ perception. It is possible that the decreased distress from the need to commute to outpatient clinics positively influenced the patients’ perception of the physician-patient relationship in our study. We hypothesize that the direct link created between physician and patient *via* the application may also give patients a feeling of exclusivity and privacy which cannot be guaranteed to the same degree in high volume outpatient clinics. Because all patients in this study had an in-person appointment at our center prior to the video call appointment, participants benefited from an established physician-patient relationship in most cases. Effects of video calls on physician-patient relationships may be different when using video calls at the first consultation. Moreover, patients in our trial consulted their physician regarding treatment-related follow-ups *via* video call. Thus, most questions concerned symptoms and side-effects and not psychologically more challenging questions such as limiting treatment at the end-of-life (16). We hypothesize that in these situations in-person interactions may have a more positive impact on the physician-patient relationship.

Our study highlights the potential of video call applications to reduce factors of socioeconomic distress in patients with solid tumors undergoing systemic cancer therapy while maintaining the physician--patient relationship. Compatibility of smartphone applications should be critically assessed prior to their implementation. Larger studies and longer follow-ups are required to assess whether differences in physical examinations, laboratory diagnostics and other factors influenced by video call consultations affect patient hospitalizations and survival.

## Data Availability

All requests for data and materials should be made to the corresponding author, following verification of any intellectual property or confidentiality obligations.

http://doi.org/10.5281/zenodo.3902837

## Ethics declarations

### Ethical approval and consent to participate

Ethical approval was granted by the University Hospital Heidelberg ethics committee on 3.4.2017 (S-090/2017). All participants provided informed consent prior to participation in the trial.

### Consent for publication

Not applicable

## Author Contributions

Conceptualization: T.W., E.C.W., J.N.K., J.K.; Questionnaire Design: J.N.K., E.C.W.; Methodology: T.W., J.N.K., E.C.W., J.K.; Data analysis: T.W, E.E., L.M.; Writing: T.W. ; Review&Editing: T.W., E.E., H.M.S., L.M., J.K., J.N.K., E.C.W.; Patient recruitment: T.W., E.E., L.M., M.S., E.G., B.G., H.H., A.I, B.K, F.K, Ath.Ma., Al. Me., A.P.M., C.F.P., D.S., H.S., N.S., L.W.; Supervision of patient care: E.W., D.J., M.D., A.S, Visualization: T.W.; Funding Acquisition: J.N.K, E.C.W., T.W.; Supervision: E.C.W., J.N.K.

